# Gonadotropin-releasing hormone antagonism reduces paedophilic interest through increased cerebellar activity

**DOI:** 10.64898/2026.04.12.26350231

**Authors:** Christian Mannfolk, Natalie Ertl, Channa N. Jayasena, Benny Liberg, Matthew B. Wall, Alexander N. Comninos, Christoffer Rahm

## Abstract

Mechanistic understanding and biomarkers of gonadotropin-releasing hormone antagonist treatment effect in paedophilic disorder are absent but may enhance outcomes and reduce sexual-offending risk. 52 help-seeking self-referred Swedish men with paedophilic disorder enrolled in a double-blinded, placebo-controlled, randomized clinical trial. Participants underwent task-based fMRI before, and two weeks after, subcutaneous injection of 120mg of degarelix or equal volume of placebo. fMRI blood-oxygen-level-dependent activation was compared between child and adult (child>adult) stimuli in task-derived regions of interest. Primary outcome was within region-of-interest child>adult activation change, whereas secondary outcomes correlated region-of-interest child>adult activation change to change in clinical measurements of risk, paedophilic interest, sexual preoccupation, hyper- and hyposexuality. 19 degarelix and 22 placebo participants had sufficient fMRI data quality. Reductions in paedophilic interest were strongly correlated with increased child>adult cerebellar (vermis) region-of-interest activation following degarelix (r=-0.740, p<0.001) but not placebo (r=0.183, p=0.41; between-group correlation coefficient z=3.347, p<0.001). Treatment did not significantly change child>adult region-of-interest activity. *Post hoc* analysis indicated that baseline autism symptoms correlated with degarelix-induced changes in paedophilic interest (r=0.717, p<0.001; between-group correlation coefficient z=2.958, p=0.003) and cerebellar activation (r=-0.581, p=0.01; between-group correlation coefficient z=-1.930, p=0.05). Increased child>adult cerebellar activation was associated with degarelix-induced reductions of paedophilic interest, suggesting cerebellar activity as mechanistically important to, and a prospective biomarker of, degarelix treatment effect. Additionally, autism symptoms may inform treatment prediction. Together, these findings have mechanistic and clinical implications for degarelix treatment of paedophilic disorder.

EU clinical trials register identifier: 2014-000647-32 https://www.clinicaltrialsregister.eu/ctr-search/trial/2014-000647-32/SE, registered on 05/06/2014.

## Introduction

Sexual interest in pre-pubertal children is the hallmark of paedophilic disorder (PeD) ^1^, and a major risk factor for committing child sexual abuse (CSA) ^2^. Despite limited clinical and mechanistic evidence, androgen deprivation therapy (ADT) is widely used to treat individuals with PeD and a high risk of sexual offending ^3–5^. ADTs are thought to decrease sexual desire through reduced androgenic signalling ^3–6^. Gonadotropin-releasing hormone (GnRH) antagonists are a subtype of ADT that inhibit luteinizing hormone and follicle-stimulating hormone secretion from the pituitary gland, drastically reducing downstream circulating androgen and oestrogen levels ^7^. GnRH antagonists have some tentative benefits over other ADTs for PeD such as faster onset of action and no initial androgen surge ^8,9^, and our recent randomized controlled trial (RCT) demonstrated that the GnRH antagonist degarelix is effective for reducing PeD symptoms and CSA risk for at least 10 weeks ^10,11^. However, the neurobiological mechanisms through which degarelix and other ADTs reduce PeD symptoms and CSA risk remains unknown ^3,4^, since no published neuroimaging studies have investigated GnRH antagonists for any indication, and only four case-reports have examined ADT in PeD with neuroimaging techniques ^12–15^.

GnRH receptors are primarily located in the pituitary gland, but also in other important brain regions for behaviour, including the olfactory bulb, hippocampus and cerebellum ^7,16^. Androgen receptors are concentrated in the limbic system, such as the hypothalamus, amygdala, and hippocampus but also present in the cerebral cortex, olfactory bulb and cerebellum ^6,17–20^. This is consistent with the limited ADT neuroimaging literature, from non-randomized studies predominantly with prostate cancer patients, that show effects on the frontal cortex ^21–23^, amygdala ^13^, hippocampus ^21^, and cerebellum ^24^.

Evaluating clinical needs, treatment effects, and CSA risk in PeD remains challenging and consequential, highlighting the need for objective, non-invasive and accurate tools ^3^. Neuroimaging, especially with relevant task-based paradigms, may enhance PeD management by identifying possible treatment targets and biomarkers ^3^.

A recent case-control study, using the Pictorial-modified Stroop task (P-MST), which measures response times to a colour tint applied over sexually salient computer-generated child and adult pictures, found increased blood-oxygen-level-dependent (BOLD) functional magnetic resonance imaging (fMRI) activation (cerebellum, and cerebral limbic and frontal structures) and response times for child compared to adult (child>adult) stimuli in individuals with PeD compared to controls ^25^. The PeD sample had significantly increased autism symptoms, a novel finding in this population, possibly affecting the observed activation pattern ^25,26^. No other studies using PeD as an inclusion criterion have examined autism; however, one recent study on child sexual abuse material consumers (60% likely had PeD) similarly reported markedly elevated autism screening scores ^27^.

Within the same project as the published PeD degarelix RCT and case-control study, we conducted a longitudinal, placebo-controlled, parallel group, double-blind, fMRI RCT to assess the effects of degarelix on BOLD signal changes during the P-MST, to explore neurobiological mechanisms and potential biomarkers.

## Methods and materials

### Study design

Participants were blinded and randomized 1:1 at the end of the baseline visit to receive either two consecutive subcutaneous injections of (a) 120mg degarelix, or (b) an equivalent volume of 0.9% sodium chloride (placebo). Participants were extensively clinically characterized by blinded assessors and underwent fMRI during performance of the P-MST prior to receiving injections (baseline) and 2 weeks after (post-intervention).

Details regarding the trial procedure have previously been published ^10^.

### Participants

Help-seeking, self-identified men with PeD were recruited between March 2016 and January 2019 through the Swedish national helpline for unwanted sexuality (PrevenTell), operated by the ANOVA sexual medicine clinic, Karolinska University Hospital, Stockholm, Sweden. Eligible participants were help-seeking men aged 18-66 years with a confirmed diagnosis of PeD, as defined by the Diagnostic Statistical Manual of Mental Disorder, Fifth edition ^1^. A full list of exclusion criteria is provided in the supplementary material or elsewhere ^10^, but is also summarized in Figure 1.

**Figure 1.**
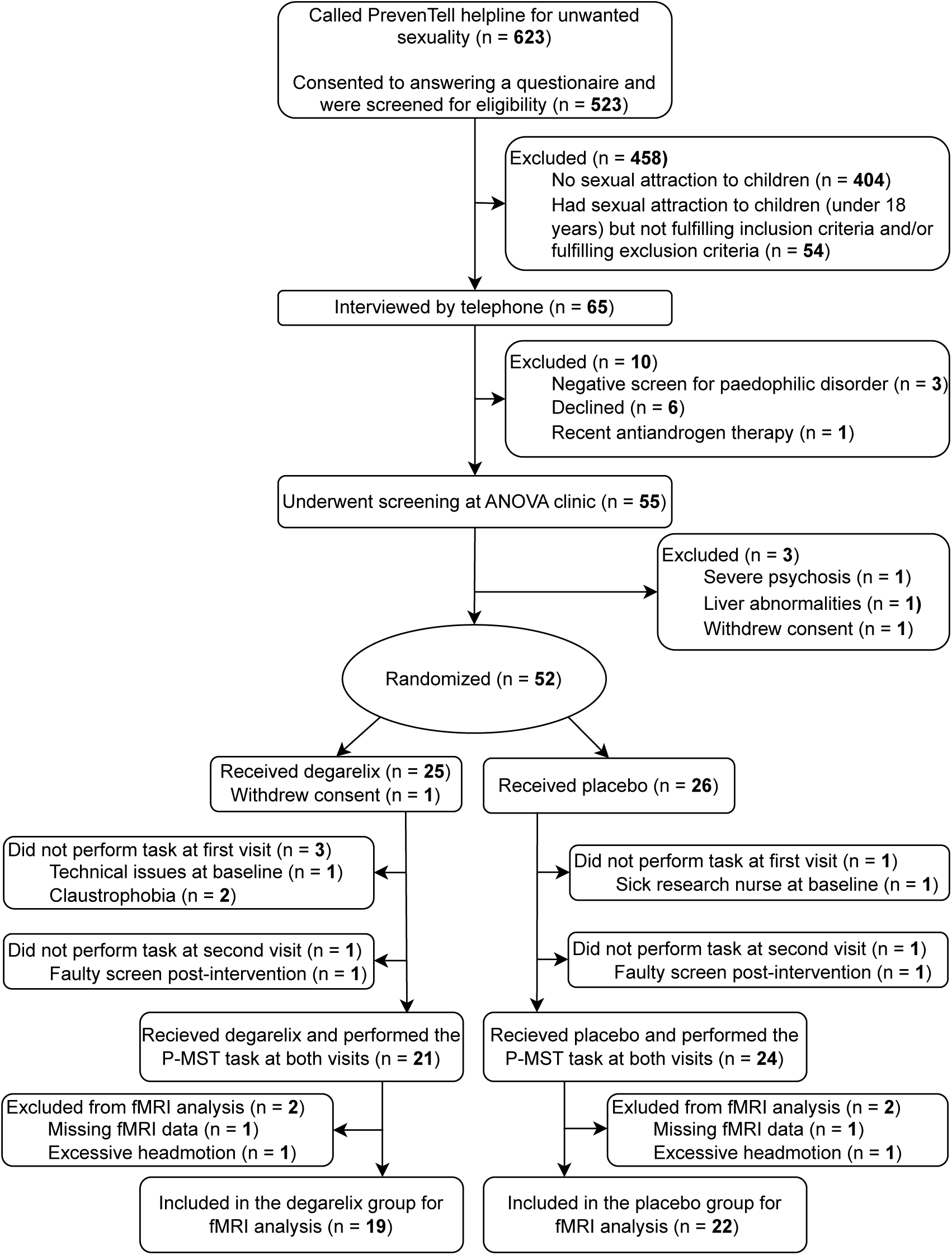
Recruitment and participant flow-chart. P-MST = Pictorial-modified Stroop task. fMRI = functional magnetic resonance imaging.

52 participants were randomized, with 19 degarelix and 22 placebo participants having fMRI data of sufficient quality and thus being included in the main fMRI analysis, for details see Figure 1 and the supplementary material.

### Clinical outcomes

A composite dynamic risk score (range: 0-15) for CSA risk was constructed for this RCT, with higher scores indicating a greater risk ^28^. It comprises five component subscales measuring: paedophilic interest, sexual preoccupation, self-regulation, empathy, and self-rated risk, each rated from 0 to 3 ^28^. Paedophilic interest was rated according to presence of PeD diagnostic criteria (i.e., presence of paedophilic attraction, and associated distress and/or behaviours) ^1^, while the other domains were evaluated using questionnaires and behavioural tests, as detailed in prior publications ^10,28^. This study used the *composite risk score* and the *paedophilic interest* and *sexual preoccupation* subscales, which previously showed significant reduction from degarelix, as compared to placebo ^10^. Hypersexuality and hyposexuality were assessed with the Hypersexuality Behaviour Inventory (HBI-19) and the Sexual Desire Inventory-2 (SDI-2), respectively ^29,30^. Autism spectrum disorder (autism) symptoms, measured with the Ritvo Autism and Asperger Diagnostic Scale-14 item screening tool (RAADS-14) ^31^, were included due to putative relevance and elevated scores in this sample ^25–27^.

### Experimental paradigm

The P-MST paradigm, originally described by Ó Ciardha ^32^, was administered as detailed in the previous P-MST case-control publication on this sample, which also includes a visualization of the paradigm ^25^. Participants performed the P-MST within the MRI scanner, indicating the colour filter applied to presented images via a four-button response pad, with instructions to respond quickly and accurately. There were two runs in succession per visit, each run consisting of 160 stimuli: 80 cat pictures (control condition), and 20 each of the four computer-generated frontal view swimwear-wearing types of human pictures from the Not Real People stimulus set: adult males (Tanner stage 5 males), adult females (Tanner stage 5 females), child males (Tanner stage 1 males), and child females (Tanner stage 1 females) ^32^. The paradigm used a block design, presenting 10 consecutive pictures from the same condition (1.5 seconds per picture), with every alternate block being a control (cat) block. Block order was pseudorandomized and differed between runs. A 30 second fixation spot preceded and followed each run. Response time and accuracy were recorded during fMRI acquisition to ensure task performance per instructions.

### Brain imaging

BOLD sensitive T2*-weighted echo planar images were collected with a Siemens Prisma 3T scanner. Each imaging volume consisted of 42 axial slices with an isotropic resolution of 3 x 3 mm (slice thickness, 3 mm; echo time, 34 ms; field of view, 240 x 240 mm; flip angle, 90°) covering the whole brain with a repetition time of 2.5 seconds. A total of 240 volumes were collected over the four runs with the first six volumes of each run discarded to allow for T1 equilibration effects. Further details of the brain imaging acquisition and processing are presented in the supplements.

### Analysis

Sample size was calculated for the published clinical outcomes ^10^ but aligns with fMRI sample sizes required to identify significant effects ^33,34^. All correlations to clinical variables were conducted using Spearman rank correlation analysis, due to modest sample size and non-parametric data (assessed by Levene’s test) in line with a previous publication ^11^.

### fMRI analysis

fMRI analysis was carried out using FMRIB Software Library (FSL) 6.0.

#### Preprocessing

Data was preprocessed following standard procedures. Data was skull-stripped, and a two-step registration process was conducted from functional to anatomical, then anatomical to standard space. The data was head-motion corrected using MCFLIRT. A high-pass temporal filter of 0.01Hz and smoothing was applied using a 6mm FWHM gaussian kernel. Participants were checked for excessive head-motion at this stage. Participants with more than 3mm of head movement in any direction were excluded from further analysis. The timings of each picture type block were convolved with the hemodynamic response function in order to create the main regressors of interest (men, women, boys, girls, cats). White matter and cerebrospinal fluid segmentations were parcellated using FMRIBs automated segmentation tool (FAST). These were co-registered to subject level functional space and thresholded at 0.5, then the mean time series was extracted, and these were used as nuisance regressors in the model,^35^ along with 24 head motion parameter, these preprocessing steps broadly followed methods used before ^36–38^. Contrasts of parameter estimates were set up to compare all contrasts of interest (adults > children, humans > cats, children > cats, men > boys etc.). Since there were two runs, a midlevel analysis was conducted in which a mean model was created of the first two runs, creating one file to be used, per subject, per visit, which was used in subsequent group level analyses.

#### ROI identification and analysis

A ROI approach was used to isolate sexual regions of the brain. To account for differences in sexual preference (regarding to teleiophilic but also gynophilic and androphilic preferences in the participants) brain areas that responded to all people (men, women, boys, girls) compared to the control cat condition were derived. ROIs were derived through constructing a group level model (all groups all visits) using the human > cat contrast. FSL cluster command (threshold Z = 5.5) was used to derive clusters. fslmaths was used to create bilateral clusters. ROI data was then extracted using fslmeants from the 4D group data, giving a value per participant, visit, and ROI. These were then used in a ROI * time (baseline – post-intervention) ANOVA per each treatment condition for the primary analysis. The primary analysis was not prespecified in the trial registration or protocol, but was deemed appropriate as a primary prior to analysis to be able to identify changes induced by degarelix treatment compared to placebo.

#### fMRI correlations

In secondary analyses, separate correlation matrices were constructed for the placebo and degarelix groups. Child>adult activation (child>adult contrast) change (post-intervention – baseline) was correlated in the ROIs with change in clinical outcomes (HBI, SDI, dynamic risk score, paedophilic interest, and sexual preoccupation) using Jamovi (version 2.3.21.0). The secondary analysis involving paedophilic interest and sexual preoccupation were prespecified in the trial protocol, published elsewhere^10^. Resulting correlations were corrected for multiple comparisons using the Benjamini-Hochberg correction, with false-discovery rate set to 0.05, k = 50.

### Exploratory analysis

fMRI correlations in the cerebellar ROI motivated *post hoc* analyses to investigate the relevance of autism spectrum symptoms, considering cerebellar differences in autism spectrum disorder ^39–42^ and elevated autism (RAADS-14) scores in this sample ^25,26^. Baseline RAADS-14 score was correlated with change in paedophilic interest and cerebellar activity. Multiple linear regression, moderation, and simple slope analyses were performed to further explore these relationships within the degarelix group, see supplements for further details.

### Ethical approval and patient consent

The National Swedish Ethics Review Appeals Board and the Swedish Medical Products Agency approved the study protocol (Ö 26-2014), and all participants provided written informed consent. The approved protocol has been published ^10^. All methodology was carried out in accordance with relevant guidelines and regulations.

### Artificial Intelligence

Artificial intelligence (ChatGPT) has only been used to improve the grammar, brevity and clarity of smaller sections of text.

## Results

### Participant characteristics

Participant characteristics are presented in Table 1.

**Table 1.** Participant characteristics at both visits. Participant characteristics of the study sample (n = 41) with sexual preference, age, and criminal convictions at baseline while the six clinical variables included in this study are presented at, and compared (within-group) between, the baseline and post-intervention visits. p<0.001 ***, p<0.01 **, p<0.05 *. SD = standard deviation; IQR = Interquartile range; HBI-19 = Hypersexual Behaviour Inventory; SDI-2 = Sexual Desire Inventory; RAADS-14 = Ritvo Autism and Asperger Diagnostic Scale – 14 item screening tool. ^a^ Missing value from one placebo participant and two degarelix participants. ^b^ Baseline and post-intervention scale values were compared with paired Wilcoxon signed-rank test, and categorical values with McNemaŕs test. ^c^ Missing value from one participant in the degarelix group at baseline. ^d^ The >13 cut-off has a 99% negative predictive value for autism spectrum disorder.^31^ ^e^ The >22 cut-off has a 37% positive predictive value for autism spectrum disorder.^31^

| Characteristics at baseline | Placebo group (n = 22) |  |  |  | Degarelix group (n = 19) |  |  |  |
| --- | --- | --- | --- | --- | --- | --- | --- | --- |
| Age in years, range, mean (SD) | 18-61, 35.64 (12.34) |  |  |  | 19-54, 34.32 (10.31) |  |  |  |
| No. (%) exclusive (only attracted to children) paedophilic disorder | 8 (36) |  |  |  | 2 (11) |  |  |  |
| No. (%) heterosexual preference for adults<sup>a</sup> | 11 (52) |  |  |  | 9 (52) |  |  |  |
| No. (%) convicted for sexual crimes. | 7 (32) |  |  |  | 4 (21) |  |  |  |
| No. (%) convicted for non-sexual crimes | 4 (18) |  |  |  | 2 (11) |  |  |  |
| Clinical outcomes, by visit | Baseline | Post-intervention | Percentual change from baseline | Change statistic<sup>b</sup> | Baseline | Post-Intervention | Percentual change from baseline | Change statistic<sup>b</sup> |
| Composite risk score (range 0-15), median (IQR)<sup>c</sup> | 8 (3) | 6.5 (4) | - 18.75 | Z = -1.83<br>p = 0.07 | 8 (3) | 5 (5) | -37.5 | Z = -3.39<br>p < 0.001<br>*** |
| Paedophilic interest (range 0-3), median (IQR) | 3 (1) | 2 (2) | -33.33 | Z = -2.55<br>p = 0.01<br>* | 2 (1) | 1 (2) | -50 | Z = -2.71<br>p = 0.007<br>** |
| Sexual preoccupation (range 0-3), median (IQR)<sup>c</sup> | 2 (1) | 2 (1) | 0 | Z = -1.34<br>p = 0.18 | 2 (1) | 1 (1) | -50 | Z = -3.09<br>p = 0.002<br>** |
| HBI-19 total sum (range 19-95), median (IQR)<sup>c</sup> | 64.5 (27) | 58.5 (33) | -9.30 | Z = -3.44<br>p < 0.001<br>*** | 54.5 (17) | 48 (25) | -11.93 | Z = -2.58<br>p = 0.01<br>** |
| No. (%) HBI-19 positive screening for hypersexuality >52<sup>c</sup> | 16 (73) | 14 (64) | -12.5 | X<sup>2</sup> = 0.50<br>p = 0.50 | 12 (67) | 5 (26) | -58.33 | X<sup>2</sup> = 4.00<br>p = 0.04<br>* |
| SDI-2 total sum (range 12-104), median (IQR)<sup>c</sup> | 74 (17) | 66 (26) | -10.81 | Z = -3.30<br>p < 0.001<br>*** | 72 (25) | 44 (33) | -38.90 | Z = -3.59<br>p < 0.001<br>*** |
| No. (%) SDI-2 positive screening for hyposexuality, <45<sup>c</sup> | 3 (14) | 4 (18) | +33.33 | X<sup>2</sup> = 0.00<br>p > 0.99 | 2 (11) | 10 (53) | +400 | X<sup>2</sup> = 5.14<br>p = 0.02<br>* |
| RAADS-14 total sum (range 0-42), median (IQR) | 14 (14) | 12.5 (17) | -10.71 | Z = -0.73<br>p = 0.46 | 15 (21) | 14 (19) | -6.67 | Z = -0.43<br>p = 0.67 |
| No. (%) RAADS-14, total score >13<sup>d</sup> | 12 (55) | 10 (46) | -20 | X<sup>2</sup> = 0.50<br>p = 0.50 | 11 (58) | 10 (53) | -9.09 | X<sup>2</sup> = 0.00<br>p > 0.99 |
| No. (%) RAADS-14, total score >22<sup>e</sup> | 5 (23) | 4 (18) | -20 | X<sup>2</sup> = 1.00<br>p > 0.99 | 5 (26) | 4 (21) | -20 | X<sup>2</sup> = 0.00<br>p > 0.99 |

### FMRI cluster identification and ROI analysis

Ten task-relevant clusters were identified and defined as ROIs (see supplementary material: Figure S1 and Table S1 for cluster details; Figure S2 for ROI visualization).

No significant differences were identified between baseline and post-intervention in the ROIs, Figure S3. To provide functional relevance for the individual participant brain changes and elucidate treatment mechanisms, change within ROI activity was correlated to change in clinical outcomes.

The correlation matrix identified a significant correlation between change (post-intervention – baseline) in paedophilic interest and change in cerebellar (centred around vermis VI, expanding into vermis crus II) ROI (child>adult contrast) activation, in the degarelix (r (17) = - 0.740, p < 0.001) but not the placebo group (r (20) = 0.183, p = 0.41), with a significant correlation coefficient difference (z = 3.347, p < 0.001), Figure 2. No other correlations between ROI activation changes and changes in clinical outcomes survived correction for multiple comparisons.

**Figure 2.**
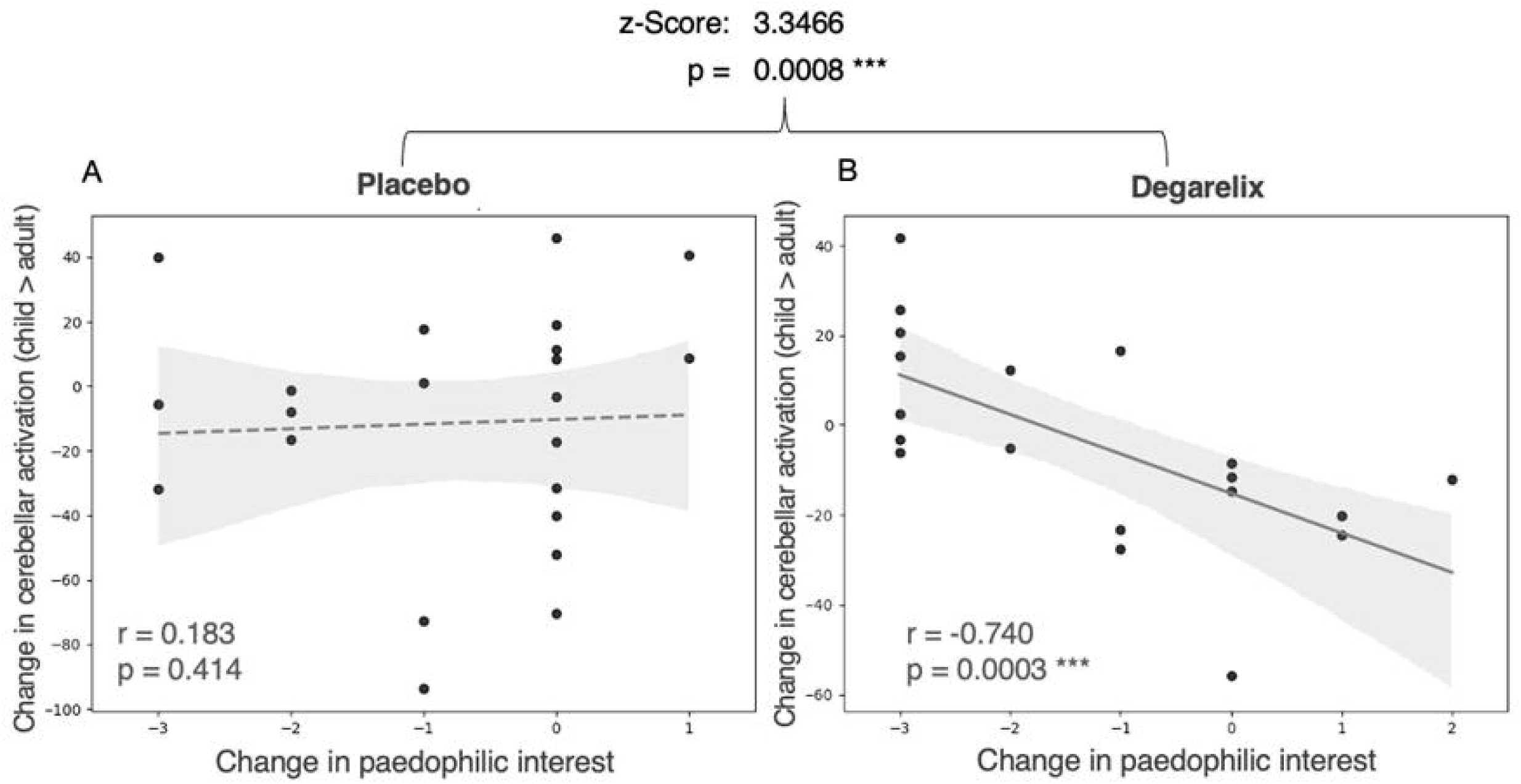
The relationship between change in paedophilic interest and change in the cerebellar blood-oxygen level-dependent activity post-intervention in the child>adult contrast, in A) the placebo group n = 22, and B) the degarelix group n = 19, Brackets between graph indicate statistical difference in the R values between the placebo and degarelix groups with each correlation, * p < 0.05, ** p < 0.01, *** p < 0.001.

### Exploratory analysis

In *post hoc* analyses, there was a significant correlation between baseline RAADS-14 autism score and change in paedophilic interest within the degarelix (r (16) = 0.717, p < 0.001) but not the placebo group (r (19) = -0.102, p = 0.66), with a significant correlation coefficient difference (z = 2.958, p = 0.003), Figure 3A.

**Figure 3.**
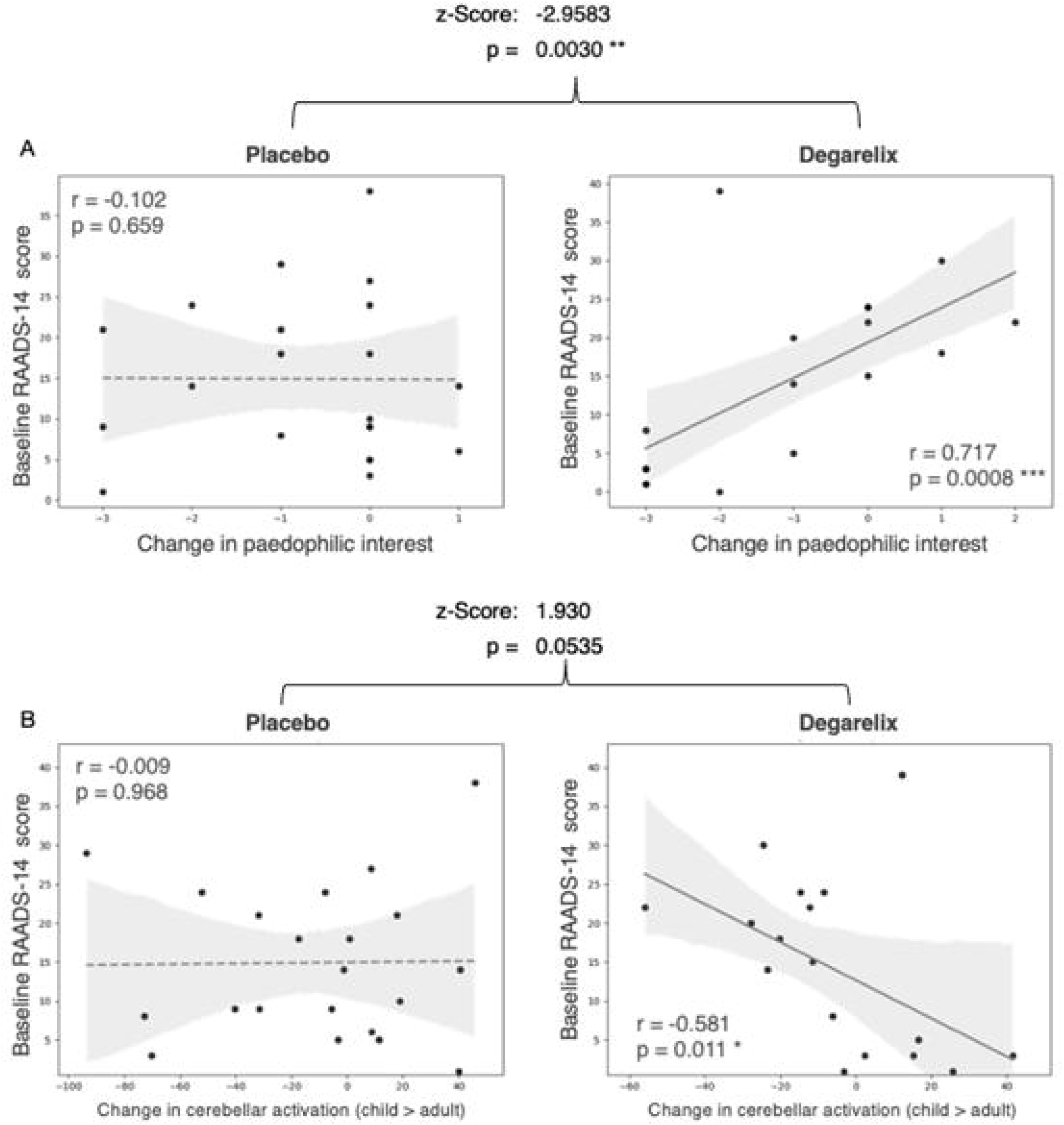
The relationship between: A) Baseline RAADS-14 autism score and change in paedophilic interest, and B) baseline RAADS-14 autism score and change in child>adult contrast cerebellar activation, in the placebo (n = 21) and degarelix (n = 18) groups. Brackets between graph indicate statistical difference in the R values between the placebo and degarelix groups with each correlation, * p < 0.05, ** p < 0.01, *** p < 0.001. RAADS-14 = Ritvo Autism and Asperger Diagnostic Scale-14 item screening tool.

There was also a significant correlation between baseline RAADS-14 autism score and change in cerebellar activity within the degarelix (r (16) = -0.581, p = 0.01) but not the placebo group (r (19) = -0.009 p = 0.97), with a non-significant correlation coefficient difference (z = -1.930, p = 0.05), Figure 3B.

A linear regression was conducted to examine whether cerebellar activation change and baseline RAADS-14 scores predicted change in paedophilic interest score from degarelix. The overall model was significant (F (2, 15) = 9.11, p = 0.003, R = 0.741, R² = 0.549) indicating that approximately 55% of the variance in change in paedophilic interest scores were explained by the predictors. RAADS-14 score was a significant predictor of change in paedophilic interest (β = 0.461, p = 0.034, whereas cerebellar activation change was not (β = -0.400, p = 0.061), when shared variance was assigned to RAADS-14.

To further probe this relationship, we conducted a simple slopes moderation analysis. The moderation interaction term was overall non-significant (Z = 3.57, p = 0.184). The simple slope analysis tests how the relationship between change in paedophilic interest and change in cerebellar activation varies over different levels of baseline RAADS-14. Average and high RAADS-14 scores were significant predictors of the relationship (B = -0.028, p = 0.022 and B = -0.046, p = 0.016) low RAADS-14 score was not (B = -0.523, p = 0.601), Figure 4.

**Figure 4.**
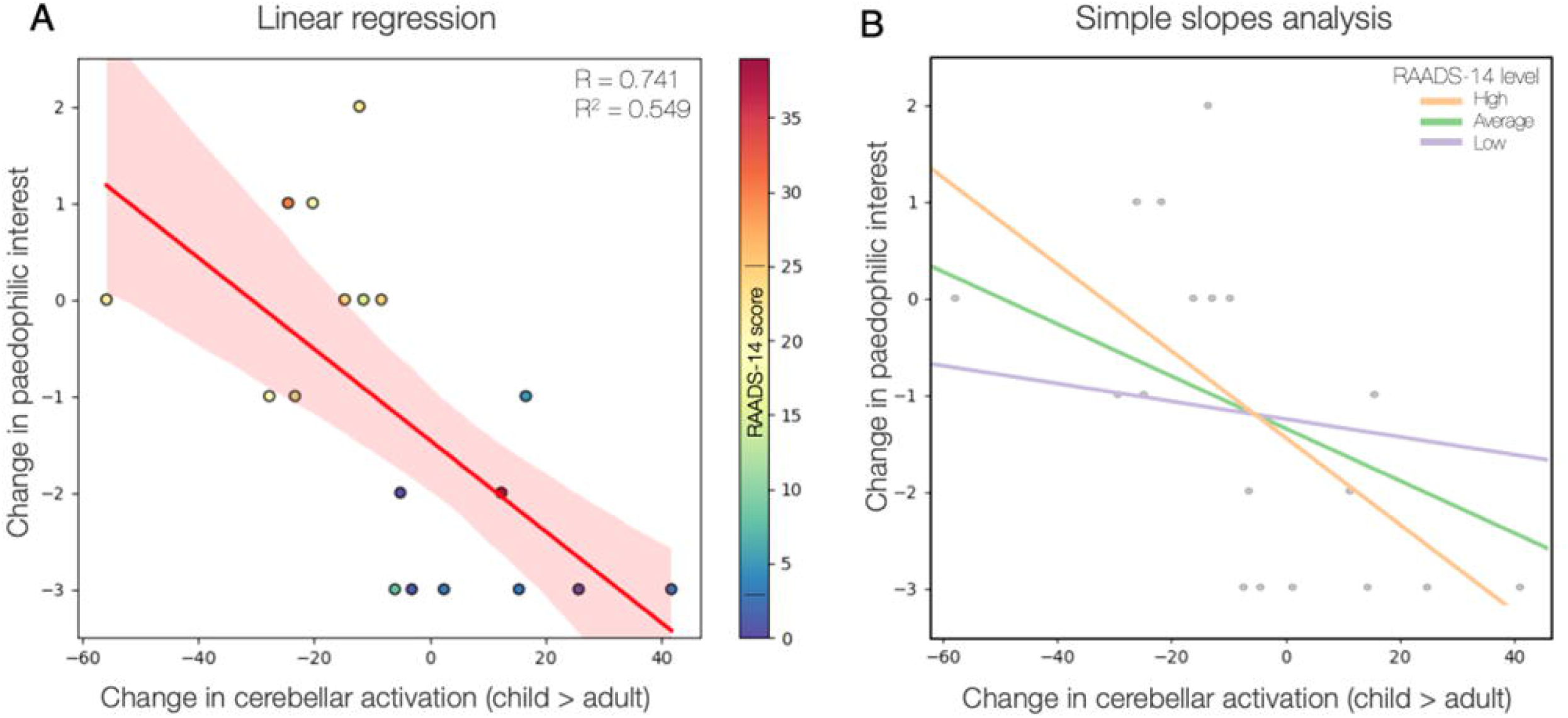
Relationship, within the degarelix group, between baseline RAADS-14 scores, degarelix-induced change in paedophilic interest and change in (child>adult contrast) cerebellar activation A) Scatterplot showing the association between change in cerebellar activation and change in paedophilic interest, with RAADS-14 scores represented by colour. B) Simple slopes analysis illustrating the interaction between cerebellar activation and RAADS-14, showing fitted regression lines for low (–1 SD), average, and high (+1 SD) RAADS-14 scores, 1SD+/- of the mean is indicated on the colour bar of figure A. N=18. RAADS-14 = Ritvo Autism and Asperger Diagnostic

## Discussion

To investigate the neurobiological mechanisms of GnRH antagonistic ADT with degarelix in PeD, this double-blinded, placebo-controlled, longitudinal fMRI study examined task-evoked activations across ten task-relevant ROIs in a paradigm designed to capture paedophilic interest, after two-weeks of degarelix treatment. A treatment-specific relationship was observed where greater degarelix-induced increases in cerebellar activity were associated with larger reductions in paedophilic interest, providing neurobiological mechanistic insight.

An additional important correlation was observed in the degarelix group between less autistic traits (measured through RAADS-14) and larger decreases in paedophilic interest. Change in child>adult cerebellar activation and autism symptoms together explained 55% of the variance in change in paedophilic interest in the linear regression, with average and above average RAADS-14 scores being better predictors.

### Association between cerebellar activity and paedophilic interest

The observed association between paedophilic interest and cerebellar activation from degarelix aligns with evidence suggesting a cerebellar role in sexual function ^43,44^ and PeD^25,45–48^ as well as social, emotional, and cognitive processing ^49–54^. Literature on cerebellar sex hormone receptors are limited and largely rodent-based ^6,17,20,55^, but suggest cerebellar sensitivity to androgens and oestrogens ^6,18,20,56^. Few human studies have investigated the male cerebellum-androgen relationship, but one study found an association between cerebellar volume and testosterone levels, suggesting that testosterone regulates negative emotions via cerebellar volume ^51^. An interesting rodent study, following previous work implying the cerebellar vermis in rodent sexual behaviour ^57,58^, identified castration-responsive androgen receptors involved in reproductive behaviour within Purkinje cells in the (posterior) rodent cerebellar vermis ^19^, with suggested relevance for the libido-lowering effect of ADT in human males ^20^.

Previous studies have demonstrated increased cerebellar activation to child>adult stimuli in men with PeD ^25^, and male paedophilic child sexual offenders ^14,46–48^. The cerebellar ROI in this study included the vermis, consistent with findings implicating the vermis in processing of salient child stimuli in men with PeD ^25,48^, ADT treatment of PeD ^14^, and child sexual offending ^45,59^. Additionally, lesions in the rodent vermis lobule VI have been shown to disrupt reproductive behaviour ^60^.

Taken together, increased cerebellar, specifically vermis, activity represents a plausible treatment target and biomarker for reduction of sexual and paedophilic interest, but could also be secondary to primary effects in other areas, such as cerebral limbic areas with high androgen receptor density ^6,17^, frequently implicated in PeD ^3,61–63^, with well-established cerebellar connections ^64,65^, especially the hypothalamus ^51,66,67^. These parts of the cerebellum, of which the vermis is central, have been called the limbic cerebellum for their involvement in affective processing ^64^, and presumably also sexual processing, due to its affective and salience-related aspects. As such, cerebellar (vermis) child>adult activation warrants further investigation as a prospective treatment biomarker for ADT in PeD.

Another consideration is that degarelix’s effect on paedophilic interest may also involve non-androgenic mechanisms, with direct luteinizing hormone, follicle stimulating hormone, oestrogen, or GnRH-signalling possibly affecting sexual outcomes. GnRH infusions into the rodent mid-brain, chicken cerebral ventricle, and GnRH-activity in the rodent olfactory bulb (similar concentration of GnRH-neurons as humans), all induce short-onset effects on reproductive behaviour, independently of the hypothalamic-pituitary-gonadal axis ^68–71^. This suggests that GnRH receptors have direct non-androgenic neural effects on sexual behaviour and desire. Interestingly, there is a high gene expression and confirmed presence of GnRH receptors in the cerebellum, with evidence for GnRH regulation of the rodent cerebellum ^7,16,72,73^. Additionally, androgen receptor blockers without direct GnRH receptor activity, such as cyproterone acetate, are considered less effective in the acute phase of PeD treatment ^4,74^. Collectively these different lines of evidence suggest that degarelix may reduce paedophilic interest partially through non-androgenic modulation of GnRH receptors, in the cerebellum (as in this study) or elsewhere. This concept holds clinical relevance, by theoretically allowing PeD treatment without necessitating castrate levels of androgens (and oestrogens), potentially enhancing treatment tolerability. Future studies should investigate the sexual relevance of extra-pituitary GnRH-receptors in humans.

### Association between baseline autism symptoms and treatment effects

Bivariate analyses indicated that lower baseline RAADS-14 scores were strongly associated with greater reduction in paedophilic interest and greater increases in child>adult cerebellar activation, only within the degarelix group. However, when both predictors were entered into a linear regression, only RAADS-14 remained a significant predictor of change in paedophilic interest, indicating the importance of autistic traits in the degarelix-induced effect. Together, RAADS-14 scores and change in cerebellar activation explained around 55% of the variance, indicating both factors are important for the treatment effect of degarelix but also that other factors, perhaps not captured in this study, may contribute to its impact on paedophilic interest.

This may reflect distinct but converging pathways, consistent with suggestions of differential cerebellar functioning in autism, including in the vermis ^39–42,64^. However, cerebellar abnormalities are common in psychiatric and neurodevelopmental pathology ^64^. Furthermore, elevated RAADS-14 scores may be related to the suggested neurodevelopmental underpinnings of PeD ^25,26,75^, and may be capturing non-specific traits related to neurodevelopmental PeD aberrations ^25,26,75^, autism-similar CSA risk factors (such as deficits in cognitive empathy or social abilities)^2,27^, or associated concepts co-occurring with paedophilic interest, such as “emotional congruence with children” ^76–78^.

Our results suggest that RAADS-14 score may inform degarelix treatment prediction, but previous RCT results from this sample, albeit with a considerably different statistical approach, show no association between baseline RAADS-14 and paedophilic interest at 10 weeks follow-up.^11^ Incorporating these differences, possible explanations include that individuals with high RAADS-14 scores are less prone, and/or have a longer latency (i.e. > 2 weeks), towards receiving and/or reassessing treatment effects, consistent with autism core features ^1^. Additionally, treatment of autistic individuals is associated with challenges related to communication, self-report, and often lower effectiveness ^79,80^. Future studies should investigate the predictive value of RAADS-14 score and proper autism diagnosis for ADT efficacy, during longer follow-up.

### Strengths and Limitations

As the first longitudinal fMRI RCT on ADT and GnRH-antagonism, that also incorporates a validated PeD-specific task paradigm, this study provides novel insights into how GnRH antagonists may reduce paedophilic interest, with implications for the broader understanding of GnRH antagonist treatment and ADT in PeD.

Several limitations are worth considering, first, the constrained and noisy fMRI environment may have impeded intended task functioning, although significant task-effects were still observed. Second, general effects of ADT^21–23^ may also lead to decreased activation to non-sexual stimuli (such as for cat or adult stimuli), thus decreasing the strength of statistical comparison within the degarelix group. Third, heterogeneity of sexual preferences may preclude significant results when combining gender categories.

## Conclusion

In conclusion, degarelix-induced reductions in paedophilic interest were predicted by increased cerebellar (primarily vermis) activation to child>adult stimuli, implicating cerebellar mechanisms in the effect of degarelix in PeD. Moreover, these degarelix-induced changes appeared partially moderated by baseline RAADS-14 autism scores. These findings constitute promising steps towards improving understanding, prediction, and evaluation of PeD treatment.

## Funding

This study was funded by grants from the Swedish Society for Medicine (Grant Nos. SLS-501421 and SLS-886481 [to CR]); the Swedish Society for Medical Research (Grant No. P14-0136 [to CR]); the Professor Bror Gadelius Foundation (20220413 [to CM]); the Söderström König Foundation (to CR); the Fredrik and Ingrid Thuring Foundation (Grant Nos. FITS-2015-00157 [to CR] and FITS-2020-00631 [to CM]); and the Centre for Psychiatry Research, Department of Clinical Neuroscience, Karolinska Institutet (Grant No. CPF-99/2016 [to CR]) and through the ALF agreement between the Swedish Government and the county councils (Grant Nos. SLL20150518 and SLL20160555 [to CR]).

## Data Availability

Data access and codes can be obtained on reasonable request.

## Acknowledgements

We thank Maria Andersson, Kerstin Eriksson, Pia Jaensen and Susanne Jarlvik Alm for help with data collection; Tomas Jonsson and Maria Kristoffersen-Wiberg for assistance with trial organization; and the staff at ANOVA and PrevenTell for contributing to participant recruitment, all at Karolinska University Hospital.

## Author Contributions

C.R. designed the study protocol and ascertained participant diagnosis. C.M processed and structured clinical data. N.E., M.W. and A.C. designed the fMRI analyses. N.E. conducted all fMRI analyses and preprocessing. N.E and C.M. conducted non-fMRI analyses and preprocessing. M.W. supervised all statistical analyses. B.L. designed the fMRI gathering and testing paradigm and was consulted for the statistical fMRI analysis plan. N.E. and C.M. created graphs, tables and wrote the primary drafts of the manuscript. All authors were involved in the development of the analysis approach, discussed results and revised the manuscript.

## Data availability

The sensitive nature of the sample and the possibility of using the data to identify the participating individuals, all whom have a highly stigmatized disorder, could put them in danger. Furthermore, the dataset could also be used to explore neuroimaging biomarkers for sexual preference, which poses serious ethical challenges and is not included in the ethical permit of this project. As such, the data will not be openly shared but can be considered for sharing with established researchers with relevant merits upon request, but may require a signed agreement regarding responsibility, non-sharing and intent.

## Conflicts of interest

N.E. and M.W. work at Perceptive which provides research consultancy services unrelated to this manuscript. The other authors declare no potential conflicts of interest.

## Notes

### Competing Interest Statement

The authors have declared no competing interest.

### Clinical Trial

EUCTR2014-000647-32

### Author Declarations

The Swedish Ethics Review Appeals Board (Oeverklagandenaemnden foer etikproevning) and the Swedish Medical Products Agency (Laekemedelsverket) gave ethical approval for this work (Oe 26-2014)

## References

1. American Psychiatric Association. Diagnostic and Statistical Manual of Mental Disorders (DSM-5®). (2013).

2. Seto, M. C., Augustyn, C., Roche, K. M. & Hilkes, G. Empirically-based dynamic risk and protective factors for sexual offending. Clin. Psychol. Rev. 106, 102355 (2023).

3. Jordan, K., Wild, T. S. N., Fromberger, P., Müller, I. & Müller, J. L. Are there any biomarkers for pedophilia and sexual child abuse? A review. Front. Psychiatry 10, 940 (2020).

4. Landgren, V. et al. Pharmacological treatment for pedophilic disorder and compulsive sexual behavior disorder: A review. Drugs 82, 663–681 (2022).

5. Culos, C., Di Grazia, M. & Meneguzzo, P. Pharmacological Interventions in Paraphilic Disorders: Systematic Review and Insights. J. Clin. Med. 13, 1524 (2024).

6. Pillerová, M. et al. On the role of sex steroids in biological functions by classical and non-classical pathways. An update. Front. Neuroendocrinol. 62, 100926 (2021).

7. Martínez-Moreno, C., Calderón-Vallejo, D., Harvey, S., Arámburo, C. & Quintanar, J. Growth Hormone (GH) and Gonadotropin-Releasing Hormone (GnRH) in the Central Nervous System: A Potential Neurological Combinatory Therapy? Int. J. Mol. Sci. 19, 375 (2018).

8. Krakowsky, Y. & Morgentaler, A. Risk of Testosterone Flare in the Era of the Saturation Model: One More Historical Myth. Eur. Urol. Focus 5, 81–89 (2019).

9. Thibaut, F. et al. The World Federation of Societies of Biological Psychiatry (WFSBP) Guidelines for the biological treatment of paraphilias. World J. Biol. Psychiatry 11, 604–655 (2010).

10. Landgren, V., Malki, K., Bottai, M., Arver, S. & Rahm, C. Effect of gonadotropin-releasing hormone antagonist on risk of committing child sexual abuse in men with pedophilic disorder: a randomized clinical trial. JAMA Psychiatry 77, 897–905 (2020).

11. Landgren, V., Olsson, P., Briken, P. & Rahm, C. Effects of testosterone suppression on desire, hypersexuality, and sexual interest in children in men with pedophilic disorder. World J. Biol. Psychiatry 23, 560–571 (2022).

12. Jordan, K., Fromberger, P., Laubinger, H., Dechent, P. & Müller, J. L. Changed processing of visual sexual stimuli under GnRH-therapy – a single case study in pedophilia using eye tracking and fMRI. BMC Psychiatry 14, 142 (2014).

13. Habermeyer, B. et al. LH-RH agonists modulate amygdala response to visual sexual stimulation: A single case fMRI study in pedophilia. Neurocase 18, 489–495 (2012).

14. Moulier, V. et al. A Pilot Study of the Effects of Gonadotropin-Releasing Hormone Agonist Therapy on Brain Activation Pattern in a Man With Pedophilia. Int. J. Offender Ther. Comp. Criminol. 56, 50–60 (2012).

15. Schiffer, B., Gizewski, E. & Kruger, T. Reduced neuronal responsiveness to visual sexual stimuli in a pedophile treated with a long-acting LH-RH agonist. J. Sex. Med. 6, 892–894 (2009).

16. Wickramasuriya, N., Hawkins, R., Atwood, C. & Butler, T. The roles of GnRH in the human central nervous system. Horm. Behav. 145, 105230 (2022).

17. Moraga-Amaro, R., Van Waarde, A., Doorduin, J. & De Vries, E. F. J. Sex steroid hormones and brain function: PET imaging as a tool for research. J. Neuroendocrinol. 30, e12565 (2018).

18. Sarkey, S., Azcoitia, I., Garcia-Segura, L. M., Garcia-Ovejero, D. & DonCarlos, L. L. Classical androgen receptors in non-classical sites in the brain. Horm. Behav. 53, 753–764 (2008).

19. Perez-Pouchoulen, M. et al. Androgen receptors in Purkinje neurons are modulated by systemic testosterone and sexual training in a region-specific manner in the male rat. Physiol. Behav. 156, 191–198 (2016).

20. Dart, D. A., Bevan, C. L., Uysal-Onganer, P. & Jiang, W. G. Analysis of androgen receptor expression and activity in the mouse brain. Sci. Rep. 14, 11115 (2024).

21. Plata-Bello, J. et al. Changes in resting-state measures of prostate cancer patients exposed to androgen deprivation therapy. Sci. Rep. 11, 23350 (2021).

22. Chao, H. H. et al. Effects of androgen deprivation on brain function in prostate cancer patients–a prospective observational cohort analysis. BMC Cancer 12, 371 (2012).

23. Chao, H. H. et al. Effects of Androgen Deprivation on Cerebral Morphometry in Prostate Cancer Patients – An Exploratory Study. PLoS ONE 8, e72032 (2013).

24. Cherrier, M. M., Cross, D. J., Higano, C. S. & Minoshima, S. Changes in cerebral metabolic activity in men undergoing androgen deprivation therapy for non-metastatic prostate cancer. Prostate Cancer Prostatic Dis. 21, 394–402 (2018).

25. Mannfolk, C., Liberg, B., Abé, C. & Rahm, C. Altered Neural and Behavioral Response to Sexually Implicit Stimuli During a Pictorial-Modified Stroop Task in Pedophilic Disorder. Biol. Psychiatry Glob. Open Sci. 3, 292–300 (2023).

26. Abé, C. et al. Brain structure and clinical profile point to neurodevelopmental factors involved in pedophilic disorder. Acta Psychiatr. Scand. 143, 363–374 (2021).

27. Lätth, J., Joleby, M., McMahan, A., Luke, T. J. & Rahm, C. Child Sexual Abuse Material Users on the Darknet: Psychiatric Morbidities Related to Offence Behavior. Sex. Abuse 10790632251347562 (2025) doi:10.1177/10790632251347562.

28. Wittström, F., Långström, N., Landgren, V. & Rahm, C. Risk Factors for Sexual Offending in Self-Referred Men With Pedophilic Disorder: A Swedish Case-Control Study. Front. Psychol. 11, 571775 (2020).

29. Spector, I. P., Carey, M. P. & Steinberg, L. The Sexual Desire Inventory: Development, factor structure, and evidence of reliability. J. Sex Marital Ther. 22, 175–190 (1996).

30. Reid, R. C., Garos, S. & Carpenter, B. N. Reliability, validity, and psychometric development of the Hypersexual Behavior Inventory in an outpatient sample of men. Sex. Addict. Compulsivity 18, 30–51 (2011).

31. Eriksson, J. M., Andersen, L. M. & Bejerot, S. RAADS-14 Screen: validity of a screening tool for autism spectrum disorder in an adult psychiatric population. Mol. Autism 4, 49 (2013).

32. Ciardha, C. Ó. & Gormley, M. Using a pictorial-modified stroop task to explore the sexual interests of sexual offenders against children. Sex. Abuse 24, 175–197 (2012).

33. Desmond, J. E. & Glover, G. H. Estimating sample size in functional MRI (fMRI) neuroimaging studies: Statistical power analyses. J. Neurosci. Methods 118, 115–128 (2002).

34. Murphy, K. & Garavan, H. An empirical investigation into the number of subjects required for an event-related fMRI study. NeuroImage 22, 879–885 (2004).

35. Power, J. D. et al. Methods to detect, characterize, and remove motion artifact in resting state fMRI. neuroimage 84, 320–341 (2014).

36. Mills, E. G. et al. Effects of Kisspeptin on Sexual Brain Processing and Penile Tumescence in Men With Hypoactive Sexual Desire Disorder: A Randomized Clinical Trial. *JAMA Netw*. Open 6, e2254313 (2023).

37. Thurston, L. et al. Effects of kisspeptin administration in women with hypoactive sexual desire disorder: a randomized clinical trial. *JAMA Netw*. Open 5, e2236131–e2236131 (2022).

38. Ertl, N. et al. Women and men with distressing low sexual desire exhibit sexually dimorphic brain processing. Sci. Rep. 14, 11051 (2024).

39. Crippa, A. et al. Cortico-cerebellar connectivity in autism spectrum disorder: what do we know so far? Front. Psychiatry 7, 20 (2016).

40. van der Heijden, M. E., Gill, J. S. & Sillitoe, R. V. Abnormal Cerebellar Development in Autism Spectrum Disorders. Dev. Neurosci. 43, 181–190 (2021).

41. Hampson, D. R. & Blatt, G. J. Autism spectrum disorders and neuropathology of the cerebellum. Front. Neurosci. 9, 420 (2015).

42. Biswas, M. S., Roy, S. K., Hasan, R. & Pk, M. M. U. The crucial role of the cerebellum in autism spectrum disorder: Neuroimaging, neurobiological, and anatomical insights. Health Sci. Rep. 7, e2233 (2024).

43. Bittoni, C. & Kiesner, J. When the brain turns on with sexual desire: fMRI findings, issues, and future directions. Sex. Med. Rev. 11, 296–311 (2023).

44. Stoléru, S., Fonteille, V., Cornélis, C., Joyal, C. & Moulier, V. Functional neuroimaging studies of sexual arousal and orgasm in healthy men and women: a review and meta-analysis. Neurosci. Biobehav. Rev. 36, 1481–1509 (2012).

45. Schiffer, B. et al. Structural brain abnormalities in the frontostriatal system and cerebellum in pedophilia. J. Psychiatr. Res. 41, 753–762 (2007).

46. Poeppl, T. B. et al. Functional cortical and subcortical abnormalities in pedophilia: a combined study using a choice reaction time task and fMRI. J. Sex. Med. 8, 1660–1674 (2011).

47. Ponseti, J. Assessment of Pedophilia Using Hemodynamic Brain Response to Sexual Stimuli. Arch. Gen. Psychiatry 69, 187 (2012).

48. Cazala, F. et al. Brain responses to pictures of children in men with pedophilic disorder: A functional magnetic resonance imaging study. Eur. Arch. Psychiatry Clin. Neurosci. 269, 713–729 (2019).

49. Van Overwalle, F., Baetens, K., Mariën, P. & Vandekerckhove, M. Social cognition and the cerebellum: A meta-analysis of over 350 fMRI studies. NeuroImage 86, 554–572 (2014).

50. Van Overwalle, F. & Mariën, P. Functional connectivity between the cerebrum and cerebellum in social cognition: A multi-study analysis. NeuroImage 124, 248–255 (2016).

51. Schutter, D. J. L. G., Meuwese, R., Bos, M. G. N., Crone, E. A. & Peper, J. S. Exploring the role of testosterone in the cerebellum link to neuroticism: From adolescence to early adulthood. Psychoneuroendocrinology 78, 203–212 (2017).

52. Magielse, N., Heuer, K., Toro, R., Schutter, D. J. L. G. & Valk, S. L. A Comparative Perspective on the Cerebello-Cerebral System and Its Link to Cognition. The Cerebellum 22, 1293–1307 (2022).

53. Wolfs, E. M. L., Klaus, J. & Schutter, D. J. L. G. Cerebellar Grey Matter Volumes in Reactive Aggression and Impulsivity in Healthy Volunteers. The Cerebellum 22, 223–233 (2022).

54. Adamaszek, M. et al. Consensus Paper: Cerebellum and Emotion. The Cerebellum 16, 552–576 (2017).

55. Dean, S. L. & McCarthy, M. M. Steroids, sex and the cerebellar cortex: implications for human disease. The Cerebellum 7, 38–47 (2008).

56. Panichi, R. et al. Inhibition of androgenic pathway impairs encoding of cerebellar-dependent motor learning in male rats. J. Comp. Neurol. 530, 2014–2032 (2022).

57. Garcia-Martinez, R. et al. Multiunit recording of the cerebellar cortex, inferior olive, and fastigial nucleus during copulation in naive and sexually experienced male rats. The cerebellum 9, 96–102 (2010).

58. Manzo, J. et al. Fos expression at the cerebellum following non-contact arousal and mating behavior in male rats. Physiol. Behav. 93, 357–363 (2008).

59. Klöckner, M. S. et al. Widespread and interrelated gray matter reductions in child sexual offenders with and without pedophilia: Evidence from a multivariate structural MRI study. Psychiatry Clin. Neurosci. 75, 331–340 (2021).

60. Ortiz-Pulido, R. et al. Sexual behavior and locomotion induced by sexual cues in male rats following lesion of Lobules VIa and VII of the cerebellar vermis. Physiol. Behav. 103, 330–335 (2011).

61. Sartori, G., Scarpazza, C., Codognotto, S. & Pietrini, P. An unusual case of acquired pedophilic behavior following compression of orbitofrontal cortex and hypothalamus by a Clivus Chordoma. J. Neurol. 263, 1454–1455 (2016).

62. Mohnke, S. et al. Brain alterations in paedophilia: a critical review. Prog. Neurobiol. 122, 1–23 (2014).

63. Storch, M. et al. Hypothalamic volume in pedophilia with or without child sexual offense. Eur. Arch. Psychiatry Clin. Neurosci. 273, 1295–1306 (2023).

64. Schmahmann, J. D., Oblak, A. L. & Blatt, G. J. Cerebellar connections with limbic circuits: anatomy and functional implications. in Handbook of the cerebellum and cerebellar disorders 1–21 (Springer, 2021).

65. Prati, J. M., Pontes-Silva, A. & Gianlorenço, A. C. L. The cerebellum and its connections to other brain structures involved in motor and non-motor functions: A comprehensive review. Behav. Brain Res. 465, 114933 (2024).

66. Haines, D. E., Dietrichs, E., Mihailoff, G. A. & McDonald, E. F. The cerebellar-hypothalamic axis: basic circuits and clinical observations. Int. Rev. Neurobiol. 41, 83–107 (1997).

67. Schutter, D. J. L. G. The cerebello-hypothalamic–pituitary–adrenal axis dysregulation hypothesis in depressive disorder. Med. Hypotheses 79, 779–783 (2012).

68. Decoster, L. et al. A GnRH neuronal population in the olfactory bulb translates socially relevant odors into reproductive behavior in male mice. Nat. Neurosci. 27, 1758–1773 (2024).

69. Dennison, E., Bain, P. A., Bartke, A. & Meliska, C. J. Systemically administered gonadotrophin-releasing hormone enhances copulatory behaviour in castrated, testosterone-treated hyperprolactinaemic male rats. Int. J. Androl. 19, 253–259 (1996).

70. Riskind, P. & Moss, R. L. Midbrain LHRH infusions enhance lordotic behavior in ovariectomized estrogen-primed rats independently of a hypothalamic responsiveness to LHRH. Brain Res. Bull. 11, 481–485 (1983).

71. Maney, D. L., Richardson, R. D. & Wingfield, J. C. Central administration of chicken gonadotropin-releasing hormone-II enhances courtship behavior in a female sparrow. Horm. Behav. 32, 11–18 (1997).

72. Albertson, A. J., Talbott, H., Wang, Q., Jensen, D. & Skinner, D. C. The gonadotropin-releasing hormone type I receptor is expressed in the mouse cerebellum. The Cerebellum 7, 379–384 (2008).

73. Skinner, D. C. et al. Effects of gonadotrophin-releasing hormone outside the hypothalamic-pituitary-reproductive axis. J. Neuroendocrinol. 21, 282–292 (2009).

74. Silvani, M., Mondaini, N. & Zucchi, A. Androgen deprivation therapy (castration therapy) and pedophilia: What’s new. Arch. Ital. Urol. E Androl. 87, 222 (2015).

75. Fazio, R. L. Toward a neurodevelopmental understanding of pedophilia. J. Sex. Med. 15, 1205–1207 (2018).

76. Fraser, J. M., Babchishin, K. M. & Helmus, L. M. Emotional Congruence with Children: An Empirical Examination of Different Models in Men with a History of Sexually Offending Against Children. Sex. Abuse 36, 546–571 (2024).

77. Abé, C. The Concept of Innate Sexual Priors in the Brain: A Theory on Why We Are Attracted to What We Are Attracted to. Sex. Cult. 29, 636–666 (2025).

78. McPhail, I. V. et al. Emotional Congruence with Children: Are Implicit and Explicit Child-Like Self-Concept and Attitude Toward Children Associated with Sexual Offending Against Children? Arch. Sex. Behav. 47, 2241–2254 (2018).

79. Weston, L., Hodgekins, J. & Langdon, P. E. Effectiveness of cognitive behavioural therapy with people who have autistic spectrum disorders: A systematic review and meta-analysis. Clin. Psychol. Rev. 49, 41–54 (2016).

80. Manter, M. A. et al. Pharmacological treatment in autism: a proposal for guidelines on common co-occurring psychiatric symptoms. BMC Med. 23, 11 (2025).

