## SupplementalMaterial for "Gonadotropin-releasing hormone antagonism reduces paedophilic interest through increased cerebellar activity"

**Supplemental material**

### Material and Methods

##### Participants and Recruitment.

Help-seeking and self-identified men were recruited through the Swedish national helpline for people experiencing unwanted or dangerous sexuality (PrevenTell), which is operated by the research centre and multidisciplinary clinic ANOVA, Karolinska University Hospital, Stockholm Sweden, which specializes in sexual medicine, andrology and transgender medicine.

Inclusion criteria were age between 18-66 years, male sex and diagnosis of paedophilic disorder as ascertained by a specialist in psychiatry (C.R. or B.L.). Exclusion criteria were degarelix contraindications (such as known osteoporosis, reduced liver function, hypersensitivity to the study drug), MRI contraindications (such a claustrophobia or intra-bodily ferromagnetic metals), impaired kidney function, ongoing drug or alcohol abuse as well as severe neurological, hormonal, or psychiatric comorbidity such as schizophrenia or brain malignities. Detailed inclusion and exclusion criteria have been published together with the study protocol, elsewhere ^1^.

52 participants were randomized to receive degarelix or placebo. One participant assigned to the degarelix group withdrew consent after randomization but before receiving the injection. Three participants in the degarelix group did not perform the P-MST at baseline (nor follow-up), two due to claustrophobia, the third due to technical errors, but likely also claustrophobia. One participant in the placebo group did not do the P-MST at baseline due to personnel sickness, and thus no one that could administer the P-MST. One participant in each group could not perform the P-MST on the second visit due to a connection error, specifically a faulty cable, to the presentation screen. Two participants from each group were excluded from analysis, one due to missing fMRI data and one due to excessive head motion in each group, for a total of four individuals excluded due to lacking fMRI data of sufficient quality.

The remaining 19 participants in the degarelix group and 22 participants in the placebo group were included in the study and the main fMRI analysis.

#### Data Gathering

The Pictorial-modified Stroop task (P-MST) and the MRI data gathering were conducted in Karolinska University Hospital, Huddinge, Sweden, with the rest of the data gathering, intervention and follow-up occurring at the ANOVA clinic.

Participants and assessors (except a study nurse that assessed adverse effects) were blinded with regards to treatment allocation, but not data analysts.

Data on sexual preferences and convictions for crimes relied on self-report. However, adult gender preferences were not asked for the three first participants in the study, explaining the missing data.

##### Clinical Outcomes

One individual in the degarelix group did not respond to any of the questionnaires at baseline (but did respond to interview questions regarding paedophilic interest) and was excluded from analyses including the Sexual Desire Inventory (SDI-2), Hypersexual Behaviour Inventory (HBI-19), dynamic risk or sexual preoccupation data. His Ritvo Autism and Asperger Diagnostic Scale – 14 item screening tool (RAADS-14) score for the baseline visit was imputed from his post-intervention visit for Table 1, since this imputed value was used in exploratory analyses as detailed below and because RAADS-14 score is not designed to change and is, alongside autistic symptoms, not expected to change between visits ^2,3^.

A cut-off for SDI-2 of a score <45 was a-priori considered as a positive screening value for hypoactive sexual desire disorder, as informed by clinical practice and in line with previous studies ^4–6^, and is thus presented in the Table 1 as a separate entry.

##### Experimental Paradigm

The P-MST was presented and responses gathered with the E-Prime v.2.0 software (Psychology Software tools, Sharpsburg, PA, USA). Participants were instructed how to perform the task using the response device, prior to entering the MRI camera. Stimuli were presented in the MRI scanner through a 32” LCD computer screen (Cambridge Research Systems, Rochester, UK) which was visible to participants through a head-coil mounted mirror. Pictures were from the Not-real-people stimulus set ^7^, as modified and provided by Ó Ciardha ^8^.

##### Brain Imaging

After functional scans had been collected, a T1-weighted anatomical image [magnetization prepared rapid acquisition gradient echo (MP-RAGE)], 176 slices; TR, 1900 ms; TE, 2.52 ms; with an isotropic voxel size of 1 mm × 1 mm × 1 mm] was acquired for all subjects. A senior consultant in neuroradiology assessed the anatomical scans of each subject for pathological signs.

Participants needed useable scans at both baseline and post-intervention to be included in the imaging analysis, due to the longitudinal nature of the analysis.

#### Analysis

There was response time and accuracy data for all participants. Task responses were checked to ensure that participants carried out the task as instructed. All individuals performed the task as instructed, except one individual in the degarelix group, that was colourblind and seemingly, albeit consistently, mixed up the response colours. Due to the consistency in his mixing up of the colours, he was included in the fMRI analysis.

Statistical analysis was done in SPSS 30 (IBM), unless otherwise specified.

##### Exploratory analysis

###### Post-hoc RAADS-14 correlations

Post-hoc bivariate correlational analysis correlated RAADS-14 to both change in paedophilic interest and child>adult cerebellar activity, within each treatment group, and the within-group correlation coefficients were statistically compared between treatment groups using Fisher Z-score.

As autism symptoms and RAADS-14 scores are not expected to change over time ^2,3^, baseline values were used, while excluding one significant outlier (greater than two standard deviations) in between visit change for each treatment group for analyses including RAADS-14 (except in Table 1). Considering having an extreme value (i.e. being an outlier) in between visit change was considered indicative of unreliable reporting in an expected static scale. For similar reasons, and as mentioned above, one individual that was missing RAADS-14 at baseline had his second visit value imputed instead.

###### Post-hoc linear regression, moderation and simple slope analysis.

Significant RAADS-14 correlations were followed up with linear regression and moderation analyses, conducted using Jamovi. The moderation model tested whether baseline RAADS-14 score moderated the relationship between change in cerebellar activation and change in paedophilic interest.

To aid interpretation of the interaction, a simple slopes analysis was performed in Jamovi. RAADS-14 scores were mean-centred, and participants were grouped into “low”, “average”, and “high” RAADS levels, corresponding to –1 SD, mean, and +1 SD from the sample mean, respectively. Predicted slopes for each group were then plotted to visualise how the strength of the relationship between cerebellar (child>adult) activation change and clinical improvement differed across levels of autistic traits.

### Results

#### fMRI Results

##### fMRI Cluster Identification and ROI Construction

We identified several clusters in the humans > cat contrast in the P-MST. See Figure S1 and Table S2 for these results. Based on these results, 10 bilateral ROIs were identified, see Figure S2 for details.

### References

1	Landgren V, Malki K, Bottai M, Arver S, Rahm C. Effect of gonadotropin-releasing hormone antagonist on risk of committing child sexual abuse in men with pedophilic disorder: a randomized clinical trial. *JAMA Psychiatry* 2020; **77**: 897–905.

2	Eriksson JM, Andersen LM, Bejerot S. RAADS-14 Screen: validity of a screening tool for autism spectrum disorder in an adult psychiatric population. *Molecular Autism* 2013; **4**: 49.

3	American Psychiatric Association. Diagnostic and Statistical Manual of Mental Disorders (DSM-5®). 2013.

4	Mannfolk C, Liberg B, Abé C, Rahm C. Altered Neural and Behavioral Response to Sexually Implicit Stimuli During a Pictorial-Modified Stroop Task in Pedophilic Disorder. *Biological Psychiatry Global Open Science* 2023; **3**: 292–300.

5	Abé C, Adebahr R, Liberg B, Mannfolk C, Lebedev A, Eriksson J *et al.* Brain structure and clinical profile point to neurodevelopmental factors involved in pedophilic disorder. *Acta Psychiatrica Scandinavica* 2021; **143**: 363–374.

6	Mills EG, Ertl N, Wall MB, Thurston L, Yang L, Suladze S *et al.* Effects of Kisspeptin on Sexual Brain Processing and Penile Tumescence in Men With Hypoactive Sexual Desire Disorder: A Randomized Clinical Trial. *JAMA Netw Open* 2023; **6**: e2254313.

7	Pacific Psychological Assesment Corporation. The NRP (Not Real People) stimulus set for assessment of sexual interest. 2004.

8	Ciardha CÓ, Gormley M. Using a pictorial-modified stroop task to explore the sexual interests of sexual offenders against children. *Sexual Abuse* 2012; **24**: 175–197.

Figure S1. Group mean (all participants both visits) brain response to pictures of people (men, women, boys, girls) compared to cats. Results are cluster corrected, Z>2.3, P<0.05

Figure S2. ROIs derived from “cluster command” for the people > cats contrast.

Figure S3. Mean change in activation to child compared to adult pictures in task-derived ROIs. Blue represents baseline and orange post-intervention, between visit change illustrated by lines connecting the dots. Placebo n = 22, degarelix n = 19.

**Table S1. Clusters activating for human but not cat stimuli.**

| Index | ROI | Voxels | *Z* MAX | X | Y | Z |
| --- | --- | --- | --- | --- | --- | --- |
| 15 | Occipital gyrus (R) | 5073 | 11.9 | 54 | -66 | 4 |
| 14 | Inferior frontal gyrus (R) | 3464 | 8.67 | 42 | 6 | 30 |
| 13 | Occipital gyrus (L) | 3350 | 11.2 | -46 | -76 | 2 |
| 12 | Cerebellum | 1137 | 10.4 | -8 | -74 | -38 |
| 11 | Inferior frontal gyrus (L) | 761 | 8.15 | -50 | 30 | 24 |
| 10 | Frontal orbital (L) | 580 | 7.64 | -28 | 12 | -18 |
| 9 | Frontal pole | 561 | 7.49 | 0 | 56 | 22 |
| 8 | Superior frontal gyrus | 489 | 7.16 | 8 | 12 | 58 |
| 7 | Parietal lobule (R) | 364 | 6.8 | 30 | -54 | 56 |
| 6 | Parietal lobule (L) | 242 | 7.38 | -30 | -58 | 54 |
| 5 | Amygdala (R) | 132 | 8.06 | 18 | -4 | -14 |
| 4 | Amygdala (L) | 131 | 8.13 | -18 | -8 | -14 |
| 3 | Inferior frontal pole | 122 | 6.78 | -4 | 48 | -14 |
| 2 | Brain stem | 45 | 6.85 | 10 | -28 | -8 |
| 1 | Cerebellum | 29 | 6.37 | -28 | -58 | -26 |

Cluster output from fsl cluster command from people > cats group result. Coordinates are in MNI152 space.
