## Supplementary figures and images for "Gonadotropin-releasing hormone antagonism reduces paedophilic interest through increased cerebellar activity"

### Figure S1

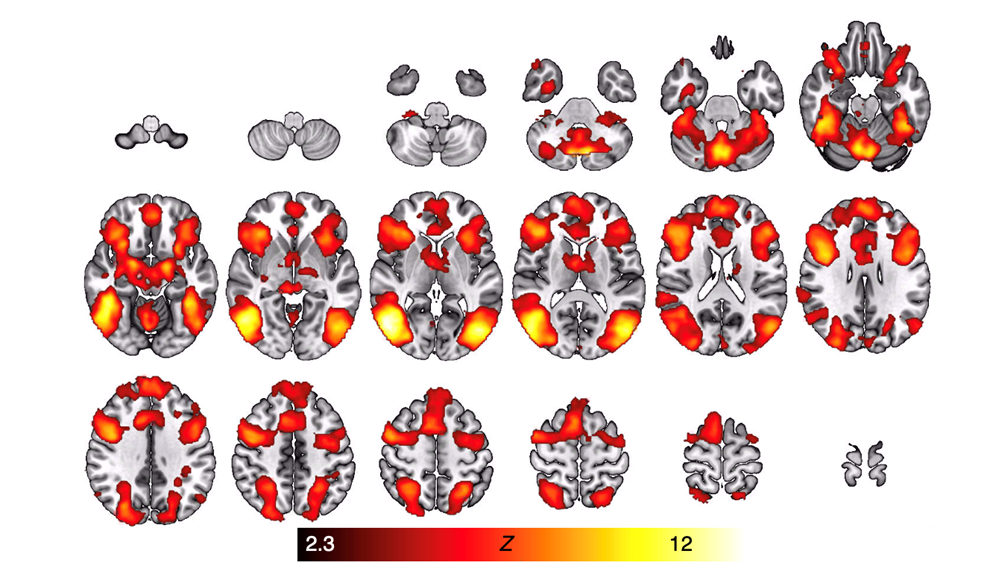

### Figure S2

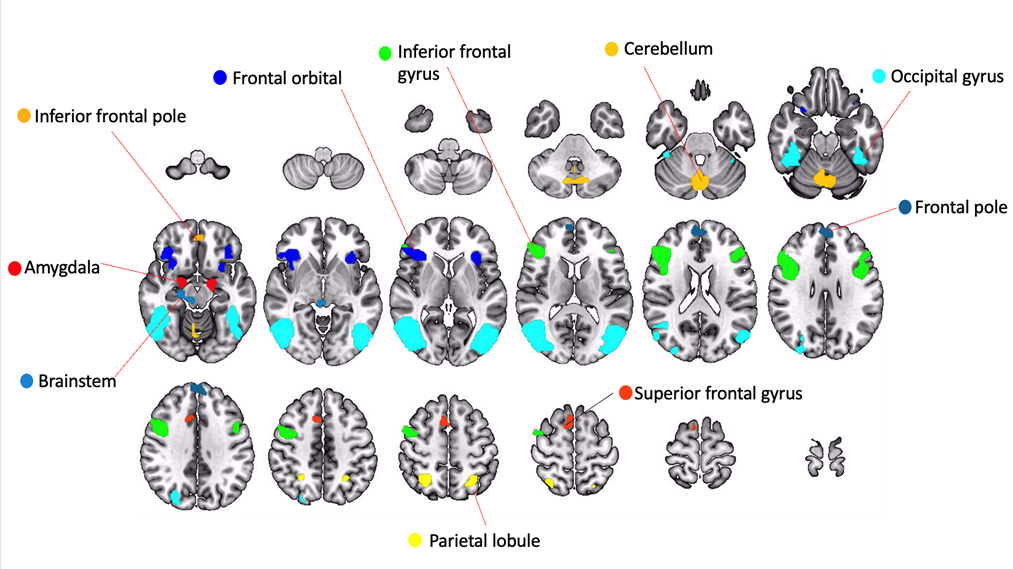

### Figure S3

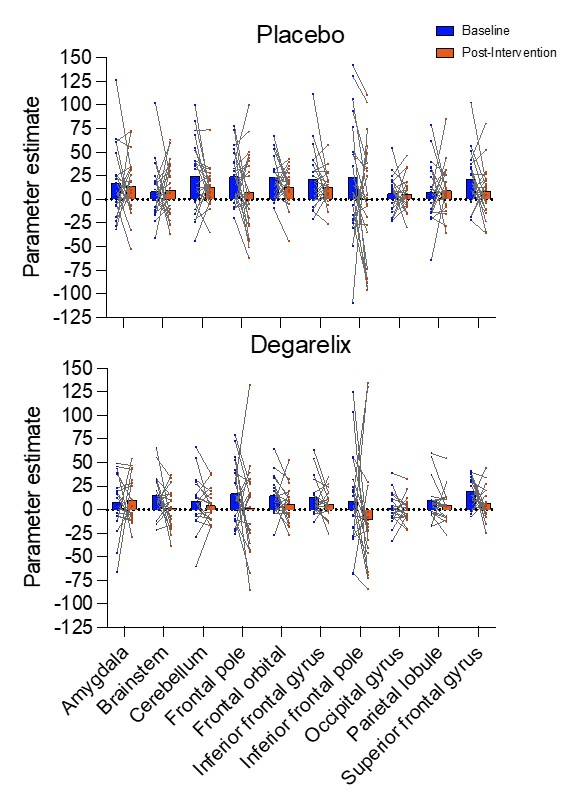
